# Computationally optimized ctDNA surveillance for recurrence detection in HPV-positive head and neck squamous cell carcinoma

**DOI:** 10.1101/2025.01.07.25320131

**Authors:** Narges Mohammadi, Ari J Rosenberg, Evgeny G Izumchenko, Alexander T Pearson, M. Reza Skandari

## Abstract

**Importance:** Early detection of Head and Neck Squamous Cell Carcinoma (HNSCC) recurrence in HPV-positive patients is crucial for improving survival rates and reducing treatment costs. Integrating circulating tumor DNA (ctDNA) testing as part of post-treatment surveillance may enhance timely cancer recurrence detection, reduce false-positive rates, and lower overall costs.

**Objective:** To develop and evaluate personalized, cost-effective post-treatment surveillance strategies that integrate ctDNA testing with established, computed tomography (CT) scans, with the goal of minimizing costs and treatment delays for HPV-positive HNSCC patients.

**Methods:** We constructed a microsimulation model that optimizes the timing of ctDNA tests and generates testing schedules designed to achieve detection delays below specified thresholds at a minimum cost. The model was fit using n= 840 training data and validated using n= 447 external data. Six sub-populations were created based on the combination of cancer stage (AJCC 8th edition stage 1, stage 2, and stage 3) and smoking status (non-smoker and ever-smoker). The study compared the proposed ctDNA-based strategy with established clinical guidelines, as well as a strategy from the literature.

**Results:** Our optimization model generated cost-effecive strategies for scheduling ctDNA tests for a range of detection delay tolerances (i.e., 3, 6, and 9 months) across the six subpopulations. The optimal ctDNA-based strategy demonstrated substantial cost savings, potentially reducing annual surveillance costs in the USA by at least $200 million compared to imaging-based guidelines, while matching an equal patient outcome of treatment delay. Additionally, a hypothetical scenario of monthly ctDNA testing, incurring comparable total cost to the existing guidelines’, offers a 32% reduction in treatment delay. The study also highlighted the growing importance of HPV-positive HNSCC surveillance, with the annual incidence projected to rise, further emphasizing the cost-saving potential of ctDNA integration.

**Conclusion:** Integrating ctDNA testing with traditional imaging methods for post-treatment surveillance of HPV-positive HNSCC patients offers a cost-effective strategy that minimizes surveillance costs and treatment delays. As the HPV-positive HNSCC population grows, the significance of the cost savings will increase. Future research should focus on the applicability of the developed strategy and their impact on patient survival and quality of life.

## Introduction

Patients with HNSCC face a significant risk of recurrence following the completion of their initial treatment. The recurrence of HNSCC is associated with a high mortality rate, with median overall survival of less than a year ^1–3^. Studies show that 80%-90% of all recurrences occur within the first two years after completion of treatment ^4, 5^. Traditional surveillance methods to monitor for recurrent cancer typically involve positron emission tomography (PET) scans and frequent computed tomography (CT) scans ^6^. However, these conventional imaging techniques present several challenges. Firstly, they are associated with high costs, which can impose a financial burden on healthcare systems and patients. Additionally, studies have shown that current imaging-based policies result in false positive results for 32% to 52% of patients, leading to a high incidence of unnecessary biopsy referrals ^7^. These frequent false positives not only drive up healthcare costs but also contribute to significant patient anxiety and stress ^8^. These limitations highlight the need for more cost-effective and optimized surveillance strategies to improve patient outcomes ^9^.

Recently, a novel assay utilizing circulating tumor-tissue DNA (ctDNA) has been developed, showing high accuracy in detecting recurrence of human papillomavirus (HPV)-positive HNSCC, which now constitutes over 70% of newly diagnosed HNSCC cases in the United States, with an annual incidence exceeding 14,000 cases ^9, 10^. The prevalence of HPV-positive HNSCC has increased dramatically, from 16% in the 1980s to 72% in the 2000s ^11^. Although various HPV assays have been evaluated, the commercially available tumor tissue modified viral (TTMV)-HPV DNA test, NavDx from Naveris, which detects circulating fragmented tumor-associated HPV DNA from apoptotic cells, has been the most extensively studied to date ^12–14^. According to Naveris data, this test is currently in clinical use at more than 400 sites ^15^. This simple blood test assay offers several advantages over traditional imaging techniques, representing a significant advancement in cancer recurrence monitoring. The benefits include lower costs, the elimination of radioactive exposure, greater convenience for patients, and lower healthcare system utilization ^9^. However, a key limitation of ctDNA is its inability to localize the recurrence site, which necessitates further imaging to determine the initiation of the treatment upon a positive ctDNA test result ^16, 17^. While studies suggest that integrating ctDNA testing with imaging can be cost-effective compared to current clinical guidelines ^8, 10^, no studies have yet developed a computationally-optimized surveillance strategy to optimize patient and system outcomes.

Studies show that early detection of recurrence can significantly enhance patient outcomes, with life expectancy gains ranging from 0.3 to 1.5 years compared to those without regular follow-up care ^18^. To minimize unnecessary strain on both patients and healthcare systems, it is crucial to address the issue of false positives results leading to unnecessary procedures. Therefore, this study aims to develop personalized, cost-effective surveillance strategies that integrate ctDNA testing with imaging, providing optimized testing schedule to improve patient outcomes and reduce healthcare costs.

## Materials and Methods

### Input parameters and recurrence pattern model parameterization

Our patient disease trajectory model is based on the model in Beesley et al. ^19^, which has been internally and externally validated and utilizes the American Joint Committee on Cancer (AJCC) 8th edition staging system. Furthermore, we conducted additional external validation of the stage-specific disease trajectories by conducting a statistical comparison test to evaluate the performance of the Beesley et al. ^19^ model against the ICON-S ^20^ model. We compared the survival probabilities generated by the ICON-S and Beesley models across three stages and over a three-year period using a *χ*^2^ test. This analysis accounted for differences in sample sizes across stages and years. The results indicated no statistically significant difference between the two models (p-value = 0.35), suggesting that both models produce comparable disease trajectories (see Figure 1 (b)).

**Figure 1:**
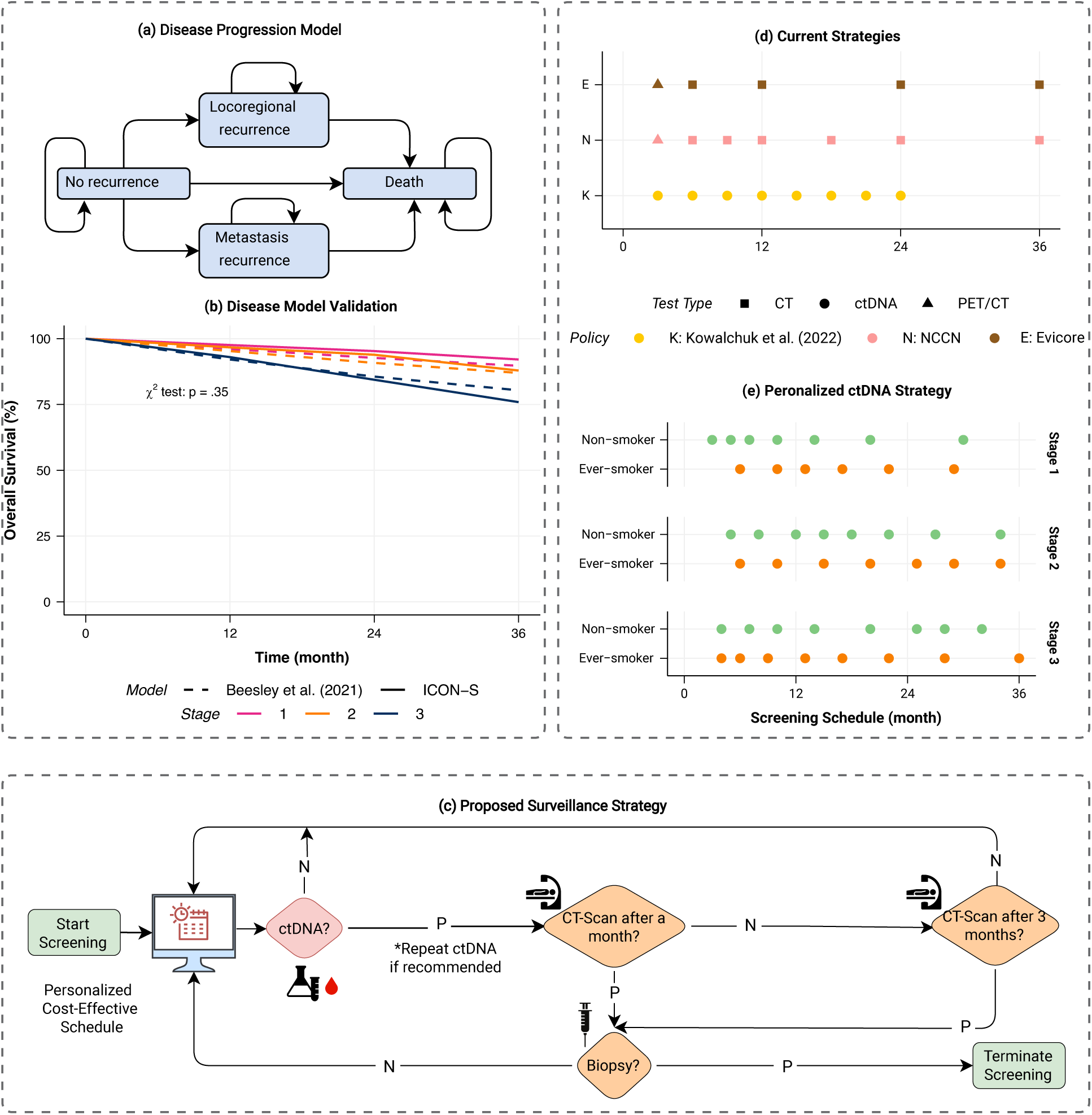
(a) Disease progression Markov model, (b) external validation of the developed model, (c) the proposed flowchart for integrating ctDNA with imaging, (d) current strategies, (e) the proposed personalized ctDNA-based schedule.

Given that majority (80% to 90%) of recurrences occur within the first two years after treatment ^4, 5^ and the original model of Beesley et al. ^19^ has a validity period of five years, our model is also limited to five years post-treatment. This approach aligns with clinical literature ^7, 8^ and guidelines ^21, 22^, which recommend surveillance for up to three years.

All model parameters including cost of imaging (including PET and CT scans), ctDNA assessments, and biopsy tests, along with their associated sensitivity and specificity values are detailed in Table 2 and Table 3. For post-treatment HNSCC, several studies report a ctDNA testing sensitivity range of 88.4% to 100%, and a specificity range of 99% to 100% ^23–27^. For these parameters, we chose the minimum reported values to ensure a conservative approach. Specifically, we used a sensitivity of 88.4%, a specificity of 99%. The cost of ctDNA testing has been reported between $500 and $1,800 ^8, 27^. For our analysis, we selected the lower end of this range ($500) to ensure consistency with the cost reference used for imaging and biopsy in comparable studies ^8^.

**Table 1:** Disease progression model parameters.

| Parameter | value | Source |
| --- | --- | --- |
| Monthly disease progression probabilities | Refer to Supplementary infomration<br>(1) | <sup>19</sup> |
| Patient covariate distribution<br>in the US population | Refer to Supplementary infomration<br>(3) | <sup>19,20</sup> |

**Table 2:** Test accuracy specifications parameters.

| Test type | Specificity | Sensitivity | Source |
| --- | --- | --- | --- |
| PET/CT-Locoregional | 87.6% | 93.7% | <sup>7</sup> |
| PET/CT-Metastasis | 93.5% | 80% | <sup>7</sup> |
| CT-Neck | 93% | 70.8% | <sup>7</sup> |
| CT-Chest | 95.4% | 45.5% | <sup>7</sup> |
| ctDNA | 99% | 88.4% | <sup>25,27</sup> |

Abbreviations:
PET: positron emission tomography;
CT: computed tomography;
ctDNA: circulating tumor-tissue;

**Table 3:**
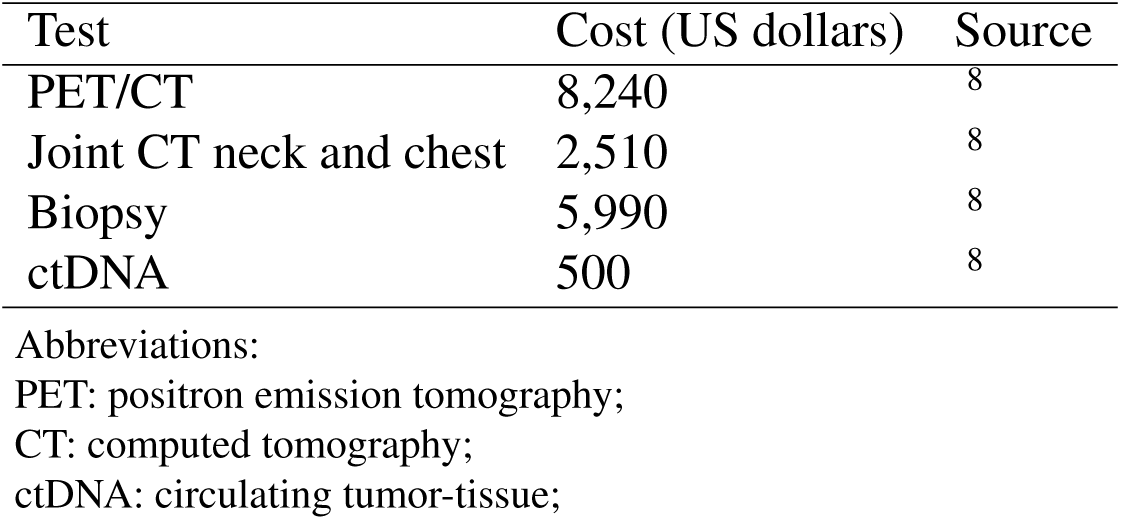
Cost parameters.

| Test | Cost (US dollars) | Source |
| --- | --- | --- |
| PET/CT | 8,240 | 8 |
| Joint CT neck and chest | 2,510 | 8 |
| Biopsy | 5,990 | 8 |
| ctDNA | 500 | 8 |

Abbreviations:
PET: positron emission tomography;
CT: computed tomography;
ctDNA: circulating tumor-tissue;

### Disease progression and simulation model

We developed a hidden Markov model to simulate post-treatment HNSCC recurrence on a monthly cycle in R (Figure 1 (a)). In this model, our focus is on symptomatic recurrence, and we assume that all patients begin in a state of no recurrence. They may transition to states representing locoregional recurrence, metastatic recurrence, or death, with transitions being time-dependent, measured relative to the time since the completion of cancer treatment. The model terminates once the patient dies or when the recurrence is detected and confirmed since the focus of the model is on the early detection phase rather than post-recurrence management. Details on the model structure and the calculation of the transition probabilities are provided in the Supplementary Information (1). To ensure robust results, we perform 20,000 simulation replications and and use the method of common random numbers to enhance precision and reduce variability ^28^. This replication number was chosen based on result convergence.

### Surveillance strategies

Several imaging-based guidelines and strategies for post-treatment surveillance of HNSCC have been proposed in the clinical literature. The National Comprehensive Cancer Network (NCCN) guideline version 1.2021 is a commonly adopted strategy, which recommends a surveillance regimen starting with a PET scan at 3 months post-treatment, followed by CT scans every three months during the first year, every six months in the second year, and a final scan at the end of the third year ^21^. The eviCore 2.1 Clinical Guidelines suggest a similar approach, initiating with a PET scan at 3 months, followed by CT scans every six months in the first year and annually thereafter (i.e., at 12, 24, and 36 months) ^22^. In both strategies, any positive imaging result is followed by a biopsy to confirm recurrence. These strategies have been depicted in Figure 1 (e).

In addition to traditional imaging-focused guidelines, emerging strategies containing ctDNA testing have been proposed as cost-effective alternatives. For example, ctDNA tests every three months for up to two years has been shown to be effective in detecting recurrences early while maintaining cost efficiency ^8^.

Our proposed strategy, detailed in the flowchart in Figure 1 (c), outlines an integrated approach for combining ctDNA testing with imaging and biopsy tests. In this strategy, any positive ctDNA result requires confirmation through joint neck and chest CT scans to localize the recurrence, followed by biopsy confirmation if imaging results are positive. Additionally, the interval between consecutive CT scans is restricted to a minimum of three months. Patient outcomes such as false positive rates and the delay in detection of recurrence as well as system outcomes such as overall cost depends on the the frequency and timing of the ctDNA tests. We have developed an optimization model that minimizes overall cost of surveillance among strategies that achieve a treatment delays below a threshold. The model is based on partially observable Markov decision process framework, which is commonly used in cancer screening and surveillance literature ^29 30^. The optimization model is described in detail in the Supplementary Information (2).

In this study, we address delays in cancer recurrence treatment caused by the lack of continuous surveillance and the imprecision of current surveillance methods. While a positive ctDNA test can indicate the presence of cancer, treatment is often delayed until the recurrence is localized through imaging and confirmed via biopsy. We define "detection delay" as the time between the actual recurrence and its detection via a positive ctDNA test. In contrast, "treatment delay" refers to the period from recurrence occurrence to its detection and eventual confirmation through imaging and biopsy, culminating in treatment initiation. Notably, "treatment delay" encompasses "detection delay". "Detection delay" is particularly relevant when comparing ctDNA-based strategies and is useful for clinicians considering initiating treatment based solely on positive ctDNA results. On the other hand, "treatment delay" provides a basis for comparing our proposed ctDNA strategy with imaging-based approaches.

### Sub-population and one-way sensitivity analysis

In this study, we present the optimal surveillance schedule for the overall population. Additionally, we categorized the population into six sub-populations based on the combination of three cancer stages (AJCC 8th Edition: stage 1, stage 2, and stage 3) of HPV-positive HNSCC and two smoking statuses (non-smoker and ever-smoker). This categorization was informed by the magnitude of hazard ratios in Beesley et al.’s model ^19^ and consultations with clinicians. Our approach aligns with prior research, including studies by Hanna et al. ^31^ and Nair et al. ^7^, which emphasize the importance of cancer stage and smoking status in post-treatment HNSCC surveillance.

To evaluate the influence of key parameters on our findings, we conduct a sensitivity analysis study focusing on the sensitivity, specificity, and cost of ctDNA testing. While the primary analysis uses the minimum reported values for these parameters reported in Table 2 and Table 3, we explore the impact of varying these values across their reported ranges on the optimal ctDNA schedule and associated outcomes. To this end, we calculate the number of ctDNA tests and the associated screening costs of achieving the target detecion delays (i.e. 3, 6, and 9 months). Specifically, we increase the sensitivity and specificity to 95% and 100%, respectively, and set the cost of ctDNA at $1800, as reported by Lin et al. ^27^, representing the maximum cost without insurance coverage.

## Results

### Optimal schedule of ctDNA test

Figure 2 illustrates the optimal schedule of ctDNA tests over a three-year period, tailored for different detection delay targets (i.e., 3, 6, and 9 months). For the population, the optimal ctDNA testing schedule includes conducting a single test at month 11 to achieve a 9-month detection delay. If a 6-month detection delay is targeted, testing should be performed at months 9, 20, and 30. To achieve a 3-month detection delay, 7 tests should be scheduled at months 5, 8, 12, 18, 21, 26, and 32. For sub-populations stratified by cancer stage and smoking status, the number and schedule of ctDNA tests differ from those of the unstratified population. Detailed information on these schedules can be found in Figure 2. Furthermore, the tabular format of the ctDNA schedule is provided in the eTable (6) in the supplementary online material. In some cases, a second ctDNA test is required to confirm an initial positive result before we advance to imaging (for example, this occurs for stage 1 non-smokers when the target detection delay is 6 months). We annotated these cases in the plot with a plus sign (+).

**Figure 2:**
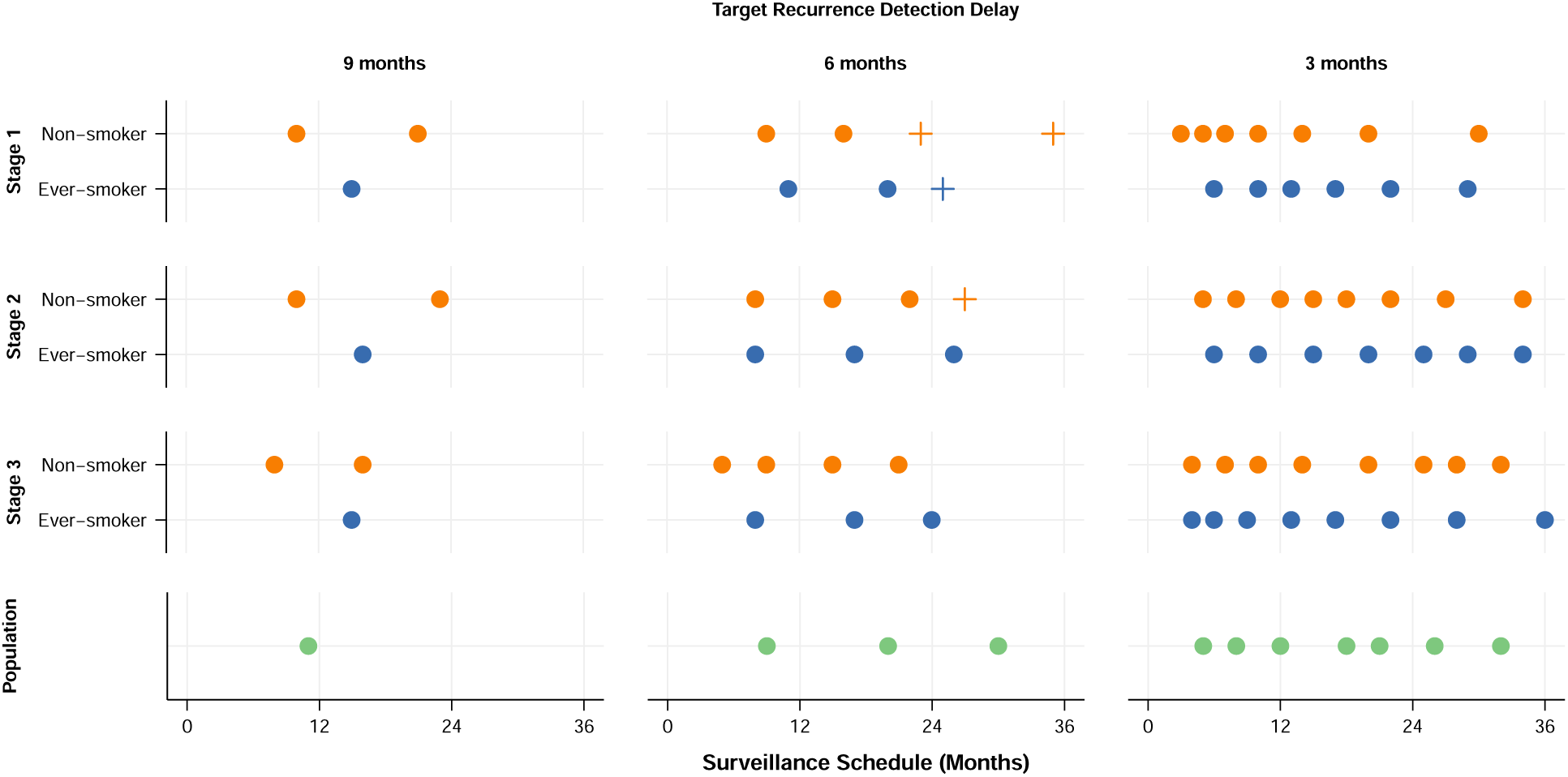
ctDNA schedule over three years for each of the six sub-populations based on three cancer stages and two smoking statuses, and unstratified population. The schedule is optimized to achieve target detection delays of 3, 6, and 9 months at the lowest cost. A positive sign (+) indicates instances where an initial positive ctDNA test requires confirmation through an additional ctDNA test before proceeding to confirmatory imaging. The tabular format of the ctDNA schedule is provided in the eTable (6) in the supplementary online material.

Figure 3 illustrates the total number of ctDNA tests and the total screening costs required to achieve the target detection delay for each sub-population. The figure clearly shows that both the number of ctDNA tests and the total cost increase as the target detection delay decreases. Additionally, the number of ctDNA tests required increases with the cancer stage.

**Figure 3:**
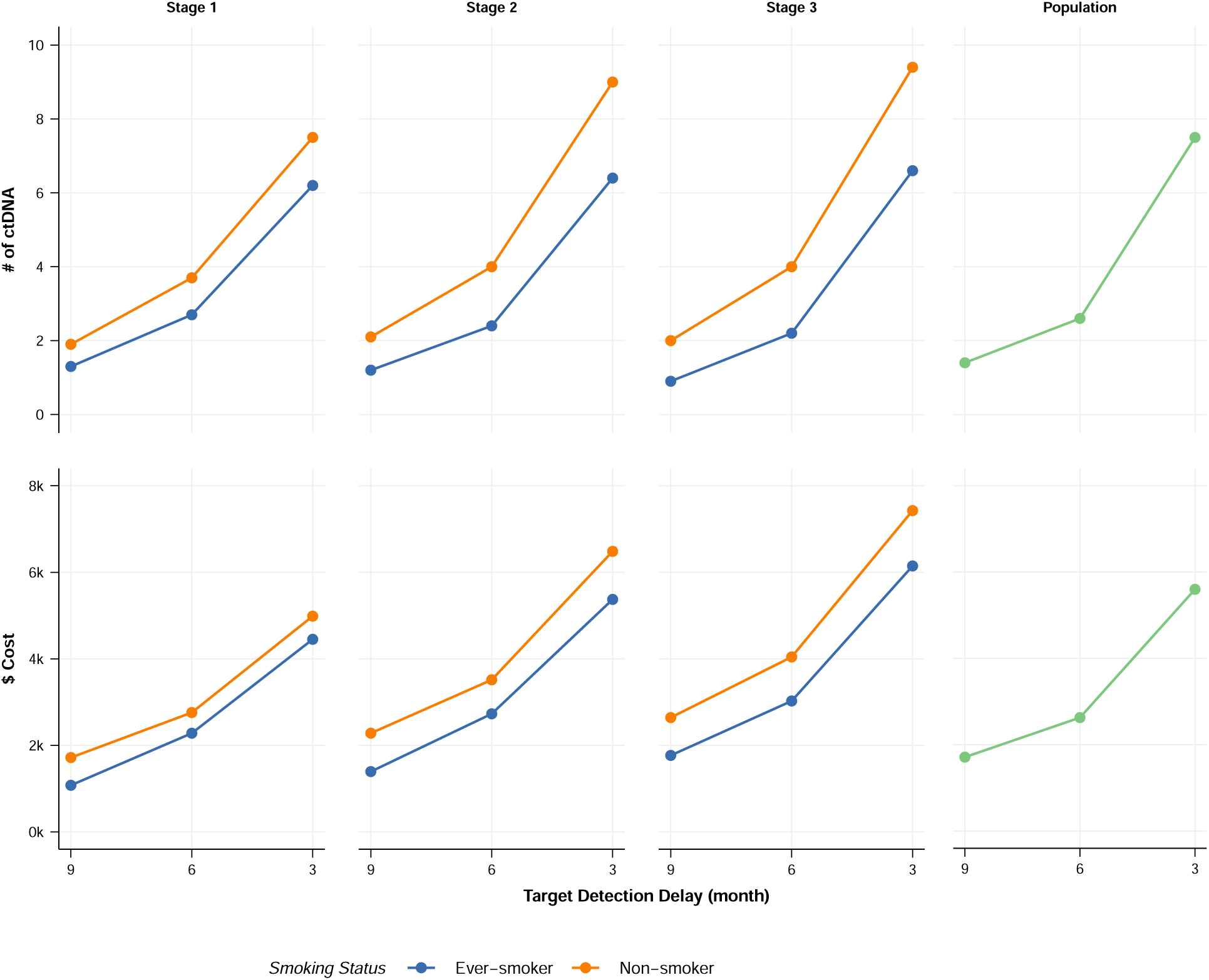
Number of ctDNA tests and total screening cost associated with each target detection delay and sub-population. The plot provides policymakers with insight into resource allocation, enabling them to adjust screening strategies according to available resources and specific population needs.

### Comparison with the existing guidelines

Figure 4 compares the schedules of current strategies with the proposed ctDNA-based strategy, and the cost savings across the U.S. population achieved by implementing ctDNA-based policies achieving the same treatment delay. Additionally, detailed comparisons of other outcomes are provided in Table 4.

**Figure 4:**
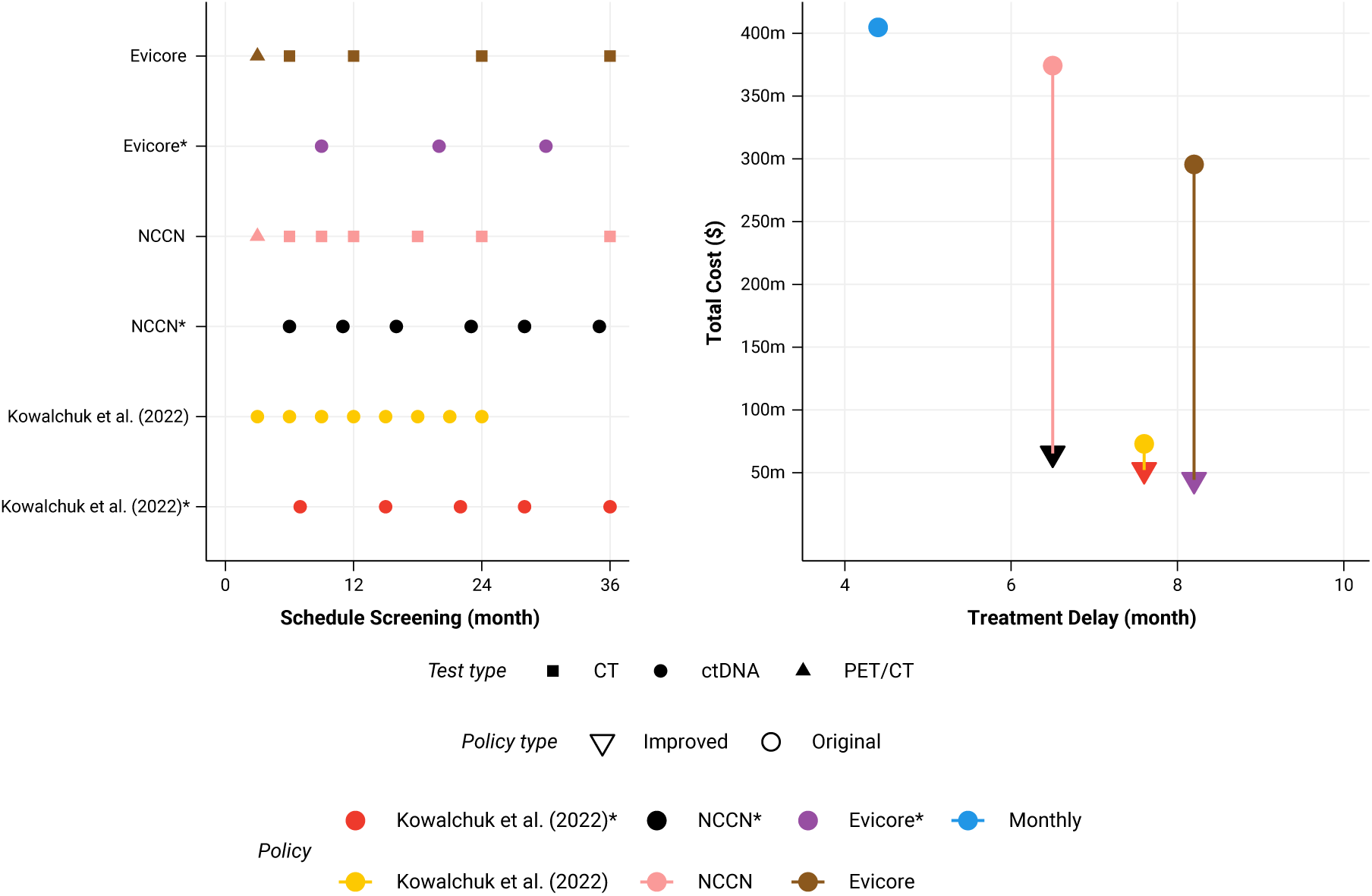
Comparison of benchmark policies with the optimal ctDNA-based strategy. This figure compares the benchmark policies (NCCN ^21^, eviCore ^22^) and the policy proposed by Kowalchuk et al. ^8^, with our proposed optimal strategy that integrates ctDNA testing and imaging, based on treatment delay and total cost for a US population of 14,000 incident HPV positive HNSCC patients. The analysis highlights the significant cost savings and clinical benefits achievable through the optimized use of ctDNA testing in conjunction with traditional imaging techniques. The strategies with * match the detection delay of the original strategy at a minimum cost through optimized ctDNA test schedule. For example, the optimal policy NCCN* achieves the same treatment delay as NCCN (6.5 months) but at a lower cost.

**Table 4:** Comparison of optimal ctDNA based policy with benchmark policies.

| Policy | Treatment delay (months) | False positive rate | Individual cost (USD) | Population cost (Million USD) | Population cost saving (Million USD) |
| --- | --- | --- | --- | --- | --- |
| NCCN | 6.5 | 56.5% | 26,731 | 374.2 | *benchmark* |
| NCCN* | 6.5 | 6.5% | 4,667 | 65.3 | 308.9 |
| EviCore | 8.2 | 46.8% | 21,110 | 295.5 | 78.7 |
| EviCore* | 8.2 | 3.7% | 3,168 | 44.4 | 329.8 |
| Kowalchuk et al. (2022) | 7.6 | 8% | 9,399 | 131.6 | 242.6 |
| Kowalchuk et al. (2022)* | 7.6 | 5.7% | 6,846 | 95.8 | 278.4 |
| Monthly test | 4.2 | 37.2% | 26,178 | 366.5 | 7.7 |

The strategies with \* match the detection delay of the original strategy at a minimum cost through optimized ctDNA test schedule. For example, the optimal policy NCCN\* achieves the same treatment delay as NCCN (6.5 months) but at a lower cost.
False positive rate: The percentage of patients ever receiving false-positive results during surveillance.
Population: U.S. incidence population, based on the annual incidence of 14,000 HPV-positive HNSCC cases.
Population cost saving: Cost savings are calculated by subtracting the cost of each strategy from the NCCN guideline's cost as the benchmark.

This analysis shows that integrating ctDNA testing with imaging can significantly reduce costs. For the annual U.S. incidence of 14,000 HPV-positive HNSCC cases, the cost savings are approximately $300 million, $250 million, and $36 million per annum compared to the NCCN, eviCore, and Kowalchuk et al. strategies, respectively ^8, 21, 22^.

The percentage of patients ever experiencing a false positive result dropped from 56.5%, 46.8%, and 8% for NCCN, eviCore, and Kowalchuk et al., down to 6.5%, 3.7%, and 5.7% for their counter ctDNA-based policies that achieve the same treatment delay. Furthermore, we found that a monthly ctDNA testing regimen, although costing nearly the same as the NCCN guideline, reduces the treatment delay by 2 months and decreases the false positive percentage in the population from 56.5% to 37.2%.

### One-way sensitivity analysis

The result of the sensitivity analysis is illustrated in Figure 5 and the result is presented for each scenario as follows:

1. Increasing sensitivity of ctDNA to 95%: By increasing the sensitivity of the test, fewer ctDNA tests are needed to reach any target detection delay, resulting in lower screening costs. The cost difference between this scenario and the baseline is approximately $100 to $300 per individual patient, and the number of ctDNA tests required decreases by 0.2 to 0.6 tests, depending on the target detection delay.
2. Increasing specificity of ctDNA to 100%: Increasing the specificity of the ctDNA test to 100% results in fewer ctDNA tests needed to reach the target detection delay. The cost difference between this scenario and the baseline is approximately $100 to $200 per individual patient, depending on the target detection delay, while the number of ctDNA tests required decreases by 0.1 tests, regardless of the target detection delay.
3. Increasing the cost of ctDNA to $1,800: We evaluated the impact of increasing the cost of ctDNA to $1,800. Our analysis indicates that, despite the increased cost, the optimal ctDNA scheduling and the total number of ctDNA tests required to achieve target detection delays of 3, 6, and 9 months remain unchanged in comparison to the baseline. However, as expected, the total screening cost increases, with the cost difference between this scenario and the baseline ranging from $1,600 to $7,600 per individual patient, depending on the target detection delay, as shown in Figure 5.

**Figure 5:**
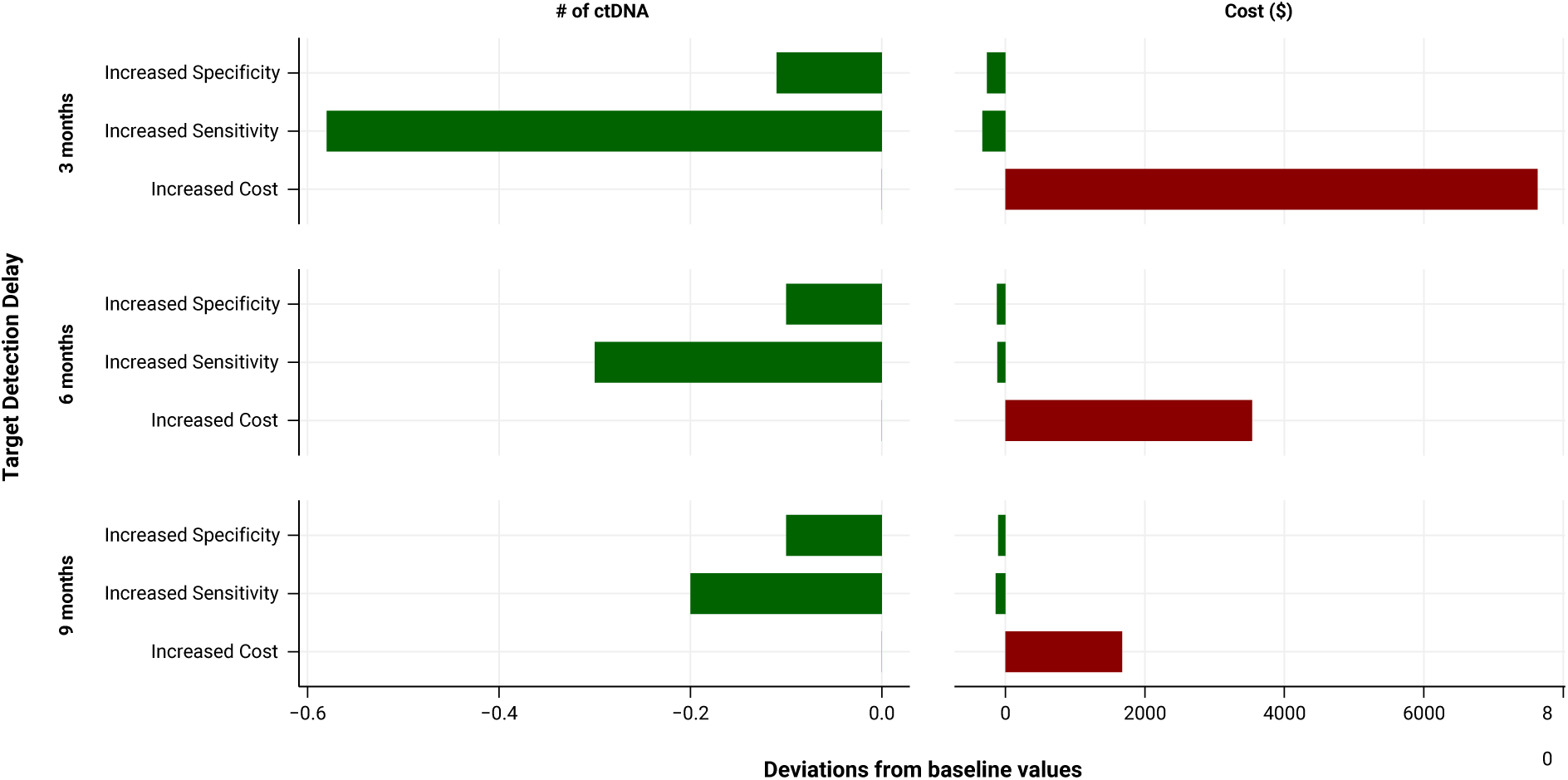
Scenario analysis results. The tornado plot illustrates the deviation from baseline values in the optimal policy performance under alternative scenarios for input parameters. The deviations are centered at zero since the baseline values differ across the three target detection delays. The bar charts are drawn relative to the base-case values, showing the sensitivity analysis for three scenarios: ctDNA’s sensitivity of 95%, specificity of 100%, and cost of $1,800.

## Discussion

The main objective of this research is to strike a balance between the total cost of screening and treatment delays, ultimately developing cost-effective surveillance strategies. Numerous clinical studies emphasize the critical importance of early detection of cancer recurrence, as it significantly impacts patient survival ^32–34^. Research indicates that early detection can lead to an increase in life expectancy by 0.3 to 1.5 months compared to those without any follow-up care ^18^. This highlights the importance of reducing treatment delay and its positive effect on patient survival achieved through surviellance. On the other hand, frequent surveillance increases costs, resource utilization, and false positive rates, which in turn lead to additional biopsy costs as well as patient anxiety and discomfort ^7, 32^. By addressing both the delay in detection and cost efficiency, our study provides a comprehensive approach to optimizing post-treatment surveillance in HNSCC patients.

Our study investigates how post-treatment surveillance can optimally detect HNSCC recurrence in HPV-positive patients by integrating ctDNA testing and CT scans. We developed a microsimulation model to evaluate alternative test schedules and strategies. We calibrated our model based on a disease trajectory model in Beesley et al. ^19^ which uses AJCC 8th edition cancer staging and has been internally and externally validated. All other input parameters are derived from clinical literature. This approach enabled us to determine the cost-effective schedule for the six sub-populations categorized by three cancer stages and two smoking statuses. Additionally, we report the results for the population without any stratification.

Our study is similar to the work by Nair et al ^7^, where a Markov model was developed to optimize surveillance regimen for the HNSCC population. Their proposed strategy is personalized based on the AJCC 7th edition cancer staging system and HPV status, focusing exclusively on optimizing CT scan schedule. They assume that a PET/CT scan is performed at three months post-treatment and then the optimal scheduling for subsequent CT scans was determined. Our results align with this study in terms of emphasizing the importance of scheduling surveillance primarily within the first three years after treatment. However, our study advances this approach by incorporating the AJCC 8th edition staging system, integrating ctDNA testing with imaging, and minimizing costs and treatment delays, thereby providing a more comprehensive and potentially more cost-effective surveillance strategy.

In this research, for all sub-populations, ctDNA screening does not commence earlier than three months after the completion of treatment. This approach aligns with clinical evidence and guidelines, which support the first post-treatment screening at approximately 12 weeks ^35^. The timing accounts for the potential dynamics of ctDNA clearance, which may vary depending on the treatment modality, such as surgery or radiotherapy (RT). Emerging evidence suggests that ctDNA levels may take time to clear following RT due to lingering tumor DNA fragments, potentially leading to false positives if screening occurs too early ^36^. This highlights a trade-off between earlier detection and the possibility of detecting residual ctDNA related to the treatment itself rather than true recurrence. As the understanding of post-treatment ctDNA dynamics evolves, it will be critical to incorporate this emerging knowledge into surveillance strategies to refine the timing of ctDNA testing. Model performance will likely change as we incorporate data for minimum residual disease testing close to the completion of treatment into future analyses.

The optimal schedule of the ctDNA test suggests that, in some cases, the ctDNA test needs to be repeated when the initial test result is positive (refer to Figure 2). This finding is of note as some clinical studies such as Chera et al ^26^ considered two consecutive abnormal ctDNA tests as a criterion to show the patient is at a higher risk of recurrence. However, we argue that conducting two consecutive tests may not always be necessary to detect recurrence, except under specific conditions. This confirmation test should generally be considered starting from the third year onward, and particularly in patients with earlier cancer stages (i.e., stage 1 and stage 2), cases where the recurrence is less likely. This more conservative approach helps mitigate the risk of false positive results, ensuring that recurrence is accurately confirmed before proceeding to further diagnostic steps.

In this research, we evaluated the guidelines including NCCN and eviCore, as well as a strategy proposed in the clinical literature, and compared them with our optimal ctDNA-based strategy. We assessed the costs associated with achieving the same treatment delay as the guidelines but through the integration of ctDNA and imaging. Our analysis demonstrates that the ctDNA-based strategy can achieve comparable treatment delays at significantly lower costs, with potential savings of $300 million and $250 million, respectively, compared to the NCCN and eviCore guidelines. Given the growing number of HPV-positive HNSCC patients ^37^, these cost savings are expected to increase over time. Additionally, the ctDNA-based strategy reduces the percentage of patients experiencing false positive results by 50% and 40% (absolute percentage reduction) when compared to the NCCN and eviCore guidelines, respectively. These cost savings are partly associated with the reduction in the percentage of patients ever experiencing false positive results, which decreases unnecessary biopsy costs. These results highlight the importance of incorporating ctDNA into surveillance programs, which not only lead to significant cost savings for the healthcare system but also reduce the emotional burden on patients due to a reduction in false positive results.

Furthermore, we show that conducting monthly ctDNA tests for up to five years incurs nearly the same cost as the NCCN guideline while resulting in a lower treatment delay (i.e., 4.2 months v.s. 6.5 months). Although performing monthly tests may not be practical, the primary aim of this analysis is to illustrate how the same expenditure as under the NCCN guideline can be utilized more effectively with a ctDNA-based strategy.

Figure 3 provides valuable guidance for policymakers, helping them adjust surveillance targets according to available resources and offering a clearer understanding of the resource implications associated with different surveillance strategies. When comparing outcomes between nonsmokers and ever-smokers within the same cancer stage, the figure indicates that ever-smokers generally require fewer ctDNA tests and incur lower screening costs. We hypothesize that this is due to the fact that smoking status does not affect the recurrence rate; rather, the lower life expectancy of ever-smokers leads to an average reduction of one ctDNA test compared to non-smokers, thereby decreasing the overall cost of screening.

One-way sensitivity analysis was conducted to evaluate the impact of uncertainty in the sensitivity, specificity, and cost of ctDNA on key outcomes, including the total cost of screening and the number of ctDNA tests required to achieve the desired detection delay targets. The analysis revealed that increasing the sensitivity or specificity of the ctDNA test reduces both the number of ctDNA tests needed and the total cost of screening, due to the corresponding decreases in false negative and false positive rates. Additionally, the scenario analysis on cost demonstrated that under a ctDNA cost increase from $500 to $1,800, the optimal ctDNA schedule and the number of ctDNA tests required to reach the target detection delay remain unchanged. However, the total screening cost increases by approximately $1,600 to $7,600 per individual, depending on the target detection delay.

This study has several limitations. First, the impact of treatment delay on more tangible patient outcomes such as life expectancy and quality of life was not investigated. Unfortunately, there is currently no comprehensive data available on the impact of surveillance on patient quality of life, necessitating future studies to collect such data for modeling assessments ^26^. Second, the primary focus of this study was on symptomatic recurrence. Although several clinical studies indicate that ctDNA is capable of detecting asymptomatic recurrence ^25^, there remains considerable debate about whether treatment should commence while the recurrence is asymptomatic and cannot be localized using imaging ^37^. Furthermore, the available clinical literature does not provide sufficient data to accurately determine disease progression probabilities for asymptomatic recurrence. Nevertheless, our proposed model can be extended to include asymptomatic conditions as relevant data becomes available. This flexibility enables future refinements and enhancements, potentially improving the model’s accuracy and applicability. Third, the focus of this study is on the HPV-positive population, and the current policies may not be optimal for the HPV-negative population. Recent clinical trials suggest that ctDNA testing could also be applicable for HPV-negative patients ^38^. However, more studies and data on the sensitivity and specificity of the test for HPV-negative patients are needed to expand the research to this population. Last, in this study, we considered screening with CT scans of the neck and chest, which are above the pelvis. However, in rare cases, the field of view of the CT scan needs to be broader. This limitation means that some patients’ recurrences may not be accurately localized with the current strategy, necessitating further investigations using additional imaging methods.

## Conclusions

In this study, we have provided personalized, cost-effective strategies for post-treatment surveillance of HPV HNSCC patients that integrates ctDNA test with traditional imaging methods. The model tailors surveillance strategy based on cancer stage and smoking status, optimizing ctDNA test schedule to achieve the same treatment delays as current guidelines while significantly reducing costs and minimizing false-positive results. As the HPV-positive HNSCC population grows, adopting such strategies will be increasingly important for both economic and patient care benefits. Moreover, the model and methods developed in this study can be adapted for other cancer types where ctDNA testing is applicable, broadening the impact across different oncological contexts. Additionally, non-invasive HPV assays have also been developed for urine and saliva ^39, 40^, which could further expand the utility of these strategies, offering additional options for patient-friendly monitoring. Future research should explore these strategies’ applicability to other populations and further examine the long-term effects of treatment delays on survival and quality of life.

## Supporting information

Supplementary Online Content

## Data Availability

All data produced in the present work are contained in the manuscript

