## Supplementary Online Content for "Computationally optimized ctDNA surveillance for recurrence detection in HPV-positive head and neck squamous cell carcinoma"

**Supplementary Online Material**

Supplementary information 1. Disease progression parameter estimation

Supplementary information 2. Optimization model and solution methodology

Supplementary information 3. Population aggregation method

eTable 1. Posterior means for baseline hazard parameters and intercepts

eTable 2. Estimated covariates associations for the multistate model extracted from Figure 2 in Beesley et al. [1]

eTable 3. Observation possibilities for screening with CT-scan

eTable 4. Observation probabilities for each observation of screening with CT-scan

eTable 5. Distribution of the patient’s characteristics in the population

eTable 6. ctDNA schedule for different target recurrence detection delay for all sub-populations

eFigure 1. Multistate model proposed by Beesley et.al [1]

eFigure 2. Markov decision process, where states 1,2, and 6 are aggregated and named “No-recurrence”

eFigure 3. Transition Probabilities over time for six sub-populations

eFigure 4. Occupancy Probabilities over time for six-sub-populations

**Supplementary information 1. Disease progression parameter estimation**

Markov and hidden Markov models are widely used in cost-effectiveness analysis, as they are more amenable to optimization compared to hidden semi-Markov models. In this study, we adapt the hidden semi-Markov model proposed by Beesley et al. [1] to develop our hidden Markov model. This adaptation preserves the occupation probabilities for all states at all times, ensuring consistency with the original model. To facilitate the optimization process, we convert the continuous-time hidden semi-Markov model into a discrete-time hidden Markov model. The adapted model, based on the framework proposed by Beesley et al. [1], is outlined in the following figure:

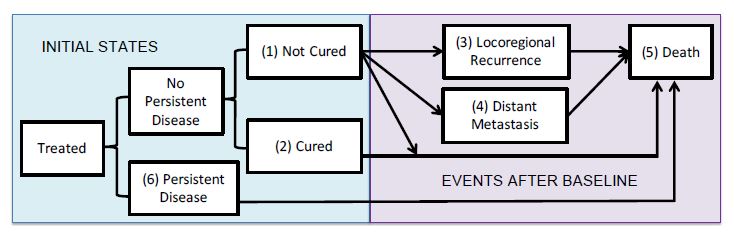

eFigure 1- Multistate model proposed by Beesley et.al [1]

This model consists of seven states of no persistent disease and not cured, no persistent disease and cured, locoregional recurrence, distant metastasis recurrence, persistent disease, and death. At time zero, the patient is in the either of not cured state with probability of $p_{1}$, cured state with probability of $p_{2}$, or in persistent disease with probability of $p_{6}$. These probabilities are estimated as follows.

**Component 1**: probability of having a persistent disease:

$$logit\left( P\left( G=6 | X \right) \right)=\gamma_{0}+\gamma_{X}X$$

$$p_{6}=P(G=6|X)$$

**Component 2**: probability of being non-cured and cured given non-persistent:

$$logit\left( P\left( G=1 | X, G\neq6 \right) \right)=\alpha_{0}+\alpha_{X}X$$

$$p_{1}=P\left( G=1 | X \right)=P\left( G=1 | X, G\neq6 \right)[1-P(G=6|X)]$$

$$p_{2}=P\left( G=2 | X \right)=\left[ 1-P\left( G=1 | X, G\neq6 \right) \right][1-P(G=6|X)]$$

Following the approach outlined in eAppendix 1 of Beesly et al. [1], which employs a proportional hazards regression model structure, in this model, $\lambda_{ij}(t)$ represents the hazard of transitioning between states $i$ and $j$ at time $t$. The covariates, denoted as $X$, include variables such as age and T-stage classifications, which are incorporated into the regression model. The coefficients associated with these covariates, $\beta_{ij}$, represent the effects of the covariates on the transition between states $i$ and $j$. These coefficients are translated into hazard ratios, capturing the relative impact of each covariate on the transition hazard. This framework allows us to estimate the parameters governing state transitions while accounting for the influence of relevant covariates.

**Component 3**: Locoregional recurrence among non-cured, non-persistent

$$\lambda_{13}\left( t \right)=\lambda_{13}^{0}\left( t \right)e^{\beta_{13}X}$$

**Component 4**: Metastasis recurrence among non-cured, non-persistent

$$\lambda_{14}\left( t \right)=\lambda_{14}^{0}\left( t \right)e^{\beta_{14}X}$$

**Component 5**: Death without prior recurrence for non-persistent patients

$$\lambda_{15}\left( t \right)=\lambda_{25}\left( t \right)=\lambda_{15}^{0}\left( t \right)e^{\beta_{15}X}$$

**Component 6**: Death for persistent patients

$$\lambda_{65}\left( t \right)=\lambda_{65}^{0}\left( t \right)$$

When modelling death rates following either locoregional recurrence or distant metastasis, the clock resets to zero upon entering the recurrence state, and the transition probabilities depend on the time spent in that state. Hence, this is a semi-Markov model. To this end, in the following formula, $T_{r}$ is the time that recurrence happens:

**Component 7**: Death after locoregional recurrence

$$\lambda_{35}\left( t-T_{r} \right)=\lambda_{35}^{0}\left( t-T_{r} \right)e^{\beta_{35}X}, t>T_{r}$$

**Component 8**: Death after metastasis recurrence

$$\lambda_{45}\left( t-T_{r} \right)=\lambda_{45}^{0}\left( t-T_{r} \right)e^{\beta_{45}X}, t>T_{r}$$

In above formula, $\lambda_{ij}^{0}(t)$ represents the baseline hazard for the transition between $i$ and $j.$

The cumulative hazard function $\Lambda_{ij}^{0}\left( t \right)=\int_{0}^{t} \lambda_{ij}^{0}\left( u \right)du$ is modelled using Weibull hazards for Components 5,7, and 8 as follows:

$$\Lambda_{ij}^{0}\left( t \right)=\left( \frac{t}{v_{ij}} \right)^{\rho_{ij}}$$

And for components 3 and 4 it follows piecewise Weibull baseline hazards as follows:

$$\Lambda_{ij}^{0}\left( t \right)=\left\{ \begin{aligned} \left( \frac{t}{v_{ij1}} \right)^{1} t\leq6 \mathrm{months} \\ \left( \frac{t}{v_{ij1}} \right)^{1}+\left( \frac{t-6}{v_{ij2}} \right)^{\rho_{ij}} t>6 \mathrm{months} \end{aligned} \right.$$

The parameters of the above model are described in eTable 1 and 2.

eTable 1 Posterior means for baseline hazard parameters and intercepts [1]

| Model: Parameter | value |
| --- | --- |
| Component 1 intercept: $\gamma_{0}$ | -4.12 |
| Component 2 intercept: $\alpha_{0}$ | -2.29 |
| Component 3 baseline hazard: $\rho_{13}$, $\log(v_{131}),$ $\log(v_{132})$ | 0.69, 4.61, 3.2 |
| Component 4 baseline hazard: $\rho_{13}$, $\log(v_{141}),$ $\log(v_{142})$ | 0.82, 5.02, 3.46 |
| Component 5 baseline hazard: $\rho_{15}$, $\log(v_{15})$ | 1.04, 7.06 |
| Component 6 baseline hazard: $\rho_{65}$, $\log(v_{65})$ | 0.68, 2.31 |
| Component 7 baseline hazard: $\rho_{35}$, $\log(v_{35})$ | 0.71, 4.21 |
| Component 8 baseline hazard: $\rho_{45}$, $\log(v_{45})$ | 0.85, 3.56 |

eTable 2 Estimated covariates associations for the multistate model extracted from Figure 2 in Beesley et al. [1]

| Covariates/ Components | C1 | C2 | C3 | C4 | C7 | C8 | C5 |
| --- | --- | --- | --- | --- | --- | --- | --- |
| Decades | -0.027 | 0.4 | -0.22 | 0 | 0.15 | -0.033 | 0.54 |
| <65 | 0 | 0 | 0 | -0.3 | 0 | 0 | 0 |
| >65 | 0 | 0 | 0 | 0.13 | 0 | 0 | 0 |
| sex: Female | 0 | -0.19 | 0.027 | -0.2 | 0 | 0 | 0.43 |
| ACE: Mild | -0.37 | -0.16 | 0 | 0 | -0.09 | -0.2 | 0.2 |
| ACE: Moderate | 0 | -0.27 | 0 | 0 | 0 | 0 | 0.54 |
| ACE: Severe | 0 | 0.46 | 0 | 0 | 0 | 0 | 0.95 |
| ACE: Moderate/Severe | 0.86 | 0 | 0 | 0 | -0.88 | 0.18 | 0 |
| Anemia: Yes | 0 | 0.8 | -0.34 | -0.4 | 0.05 | 0.48 | 0.77 |
| Smoking: Current | 0 | 0.7 | 0 | 0 | 1.4 | 0.47 | 0.9 |
| Smoking: Former | 0 | 0.3 | 0 | 0 | 0.25 | 0.73 | 0.26 |
| p16: negative | 1.3 | 0.4 | 0.7 | -0.4 | 0.57 | 0.6 | 0.76 |
| CT: 2 | 0 | 0.33 | -0.2 | 0.18 | 0 | 0 | 0 |
| CT: 3 | 0 | 0.85 | -0.33 | -0.21 | 0 | 0 | 0 |
| CT: 4 | 1.4 | 1.26 | -1.2 | -0.046 | 0 | 0 | 0 |
| CN: 2 | 0 | 0.52 | -0.22 | 0.16 | 0 | 0 | 0 |
| CN: 3 | 0 | 1.05 | -0.56 | 0.74 | 0 | 0 | 0 |

Following their model, we provide the below formula for calculating the transition probabilities for the discrete time Markov model, where state jumps happen at discrete time epochs.

- For $t=1$:

The probability of staying at state (1) at t=1 satisfies the fact that the patient is in state (1) with probability $p_{1}$ and remains there until the end of t=1.

$$\Pr_{1} \left\{ 1 | 1 \right\}=p_{1}.exp\{-Ʌ_{13}\left( t \right)-Ʌ_{14}\left( t \right)-Ʌ_{15}\left( t \right)\}$$

The probability of transition from state (1) to state (3) at time t=1 reflects the fact that the patient is in state (1) with probability $p_{1},$ and the transition to state (3) occurs at any time between t=0 and t=1, provided no death event occurs.

131=𝑝1.01exp−Ʌ13𝑣−Ʌ14𝑣−Ʌ15𝑣.𝜆13𝑣.exp−Λ351−𝑣𝑑𝑣$\Pr_{1} \left\{ 3 | 1 \right\}=p1.\int_{0}^{1} \exp\left\{ -Ʌ_{13}\left( v \right)-Ʌ_{14}\left( v \right)-Ʌ_{15}\left( v \right) \right\}.\lambda_{13}\left( v \right).\exp\left\{ -\Lambda_{35}\left( 1-v \right) \right\}dv$

The probability of transition from state (1) to state (4) at time t=1 satisfies the following equation:

141=𝑝1.01exp−Ʌ13𝑣−Ʌ14𝑣−Ʌ15𝑣.𝜆14𝑣.exp−Λ451−𝑣𝑑𝑣$\Pr_{1} \left\{ 4 | 1 \right\}=p1.\int_{0}^{1} \exp\left\{ -Ʌ_{13}\left( v \right)-Ʌ_{14}\left( v \right)-Ʌ_{15}\left( v \right) \right\}.\lambda_{14}\left( v \right).\exp\left\{ -\Lambda_{45}\left( 1-v \right) \right\}dv$

The probability of transition from state (1) to state (5) at time t=1 accounts for the fact that the patient is in state (1) with probability $p_{1}$, and death may occur at any time between t=0 and t=1. During this interval, the patient might experience locoregional or metastatic recurrence and die due to these reasons, or die without transitioning through these states.

$$\Pr_{1} \left\{ 5 | 1 \right\}=p1.\int_{0}^{1} \exp\left\{ -Ʌ_{13}\left( v \right)-Ʌ_{14}\left( v \right)-Ʌ_{15}\left( v \right) \right\}.\lambda_{15}\left( v \right)dv+$$

$$p1.\int_{0}^{1} \exp\left\{ -Ʌ_{13}\left( v \right)-Ʌ_{14}\left( v \right)-Ʌ_{15}\left( v \right) \right\}.\lambda_{13}\left( v \right).[1-exp \left\{ -\Lambda_{35}\left( 1-v \right) \right\}]dv+$$

$$p1.\int_{0}^{1} \exp\left\{ -Ʌ_{13}\left( v \right)-Ʌ_{14}\left( v \right)-Ʌ_{15}\left( v \right) \right\}.\lambda_{14}\left( v \right).[1-exp \left\{ -\Lambda_{45}\left( 1-v \right) \right\}]dv$$

The probability of staying at state (3) at t=1 reflects the fact that the patient does not experience death before t=1 and stays in state (3) throughout this time.

$$\Pr_{1} \left\{ 3 | 3 \right\}=exp\{-\Lambda_{35}\left( t \right)\}$$

The probability of transition from (3) to state (5) at t=1 is as follows:

$$\Pr_{1} \left\{ 5 | 3 \right\}=1-\Pr_{1} \{3|3\}$$

The probability of staying at state (4) at t=1 is as follows:

$$\Pr_{1} \left\{ 4 | 4 \right\}=exp\{-\Lambda_{45}\left( t \right)\}$$

The probability of transition from state (4) to state (5) at t=1 is as follows:

$$\Pr_{1} \left\{ 5 | 4 \right\}=1-\Pr_{1} \left\{ 4 | 4 \right\}$$

The probability of staying at state (2) at t=1 reflects the fact the patient is at state (2) with probability $p_{2}$ and does not experience death until time t=1.

$$\Pr_{1} \left\{ 2 | 2 \right\}=p_{2}exp\{-\Lambda_{25}\left( t \right)\}$$

The probability of transition from state (2) to state (5) at t=1 is as follows:

$$\Pr_{1} \left\{ 2 | 2 \right\}=p_{2}\left[ 1-\exp\left\{ -\Lambda_{25}\left( t \right) \right\} \right]$$

The probability of staying at state (6) at t=1 reflects the fact the patient is at state (6) with probability $p_{6}$ and does not experience death until time t=1.

$$\Pr_{1} \left\{ 6 | 6 \right\}=p_{6}exp\{-\Lambda_{65}\left( t \right)\}$$

The probability of transition from state (6) to state (5) at t=1 is as follows:

$$\Pr_{1} \left\{ 6 | 6 \right\}=p_{6}\left[ 1-\exp\left\{ -\Lambda_{65}\left( t \right) \right\} \right]$$

- For $t>1$:

The transition probability from state (1) to state (3) represents the conditional probability of transitioning to state (3) between time t and t+1, given that the patient remains in state (1) until time t. The numerator represents the probability of transitioning from state (1) to state (3) at any time between t and t+1 without a death event occurring, while the denominator represents the probability of the patient remaining in state (1) until time t.

$$\Pr_{t} \left\{ 3 | 1 \right\}=\frac{\int_{t}^{t+1} \exp\left\{ -Ʌ_{13}\left( v \right)-Ʌ_{14}\left( v \right)-Ʌ_{15}\left( v \right) \right\}.\lambda_{13}\left( v \right).\exp\left\{ -\Lambda_{35}\left( t+1-v \right) \right\}dv}{exp\{-Ʌ_{13}\left( t \right)-Ʌ_{14}\left( t \right)-Ʌ_{15}\left( t \right)\}}$$

The transition probability from state (1) to state (4) represents the conditional probability of transitioning to state (4) between time t and t+1, given that the patient remains in state (1) until time t.

$$\Pr_{t} \left\{ 4 | 1 \right\}=\frac{\int_{t}^{t+1} \exp\left\{ -Ʌ_{13}\left( v \right)-Ʌ_{14}\left( v \right)-Ʌ_{15}\left( v \right) \right\}.\lambda_{14}\left( v \right).\exp\left\{ -\Lambda_{45}\left( t+1-v \right) \right\}dv}{exp\{-Ʌ_{13}\left( t \right)-Ʌ_{14}\left( t \right)-Ʌ_{15}\left( t \right)\}}$$

The transition probability from state (1) to state (5) represents the conditional probability of transitioning to state (5) between time t and t+1, given that the patient remains in state (1) until time t. The numerator accounts for the probability that the patient is in state (1) and dies at some point between t and t+1. During this interval, the patient may experience locoregional or metastatic recurrence and die due to these reasons or die without transitioning through these states. The denominator represents the probability that the patient remains in state (1) until time t.

$$\Pr_{t} \left\{ 5 | 1 \right\}=\left[ \frac{1}{\exp\left\{ -Ʌ_{13}\left( t \right)-Ʌ_{14}\left( t \right)-Ʌ_{15}\left( t \right) \right\}} \right]\left[ \int_{t}^{t+1} \exp\left\{ -Ʌ_{13}\left( v \right)-Ʌ_{14}\left( v \right)-Ʌ_{15}\left( v \right) \right\}.\lambda_{15}\left( v \right)dv +\int_{t}^{t+1} \exp\left\{ -Ʌ_{13}\left( v \right)-Ʌ_{14}\left( v \right)-Ʌ_{15}\left( v \right) \right\}.\lambda_{13}\left( v \right).[1-exp \left\{ -\Lambda_{35}\left( t+1-v \right) \right\}]dv+\int_{t}^{t+1} \exp\left\{ -Ʌ_{13}\left( v \right)-Ʌ_{14}\left( v \right)-Ʌ_{15}\left( v \right) \right\}.\lambda_{14}\left( v \right).[1-exp \left\{ -\Lambda_{45}\left( t+1-v \right) \right\}]dv \right]$$

The probability of remaining in state (1) represents the conditional probability of staying in state (1) between time t and t+1, given that the patient is in state (1) and alive at time t.

$$\Pr_{t} \left\{ 1 | 1 \right\}=\frac{exp\{-Ʌ_{13}\left( t+1 \right)-Ʌ_{14}\left( t+1 \right)-Ʌ_{15}\left( t+1 \right)\}}{exp\{-Ʌ_{13}\left( t \right)-Ʌ_{14}\left( t \right)-Ʌ_{15}\left( t \right)\}}$$

The probability of remaining in state (3) represents the conditional probability of staying in state (3) between time t and t+1 and death does not happen, given that the patient is in state (3) and alive at time t.

$$\Pr_{t} \left\{ 3 | 3 \right\}=\frac{\int_{0}^{t} \exp\left\{ -Ʌ_{13}\left( v \right)-Ʌ_{14}\left( v \right)-Ʌ_{15}\left( v \right) \right\}.\lambda_{13}\left( v \right).[exp \left\{ -\Lambda_{35}\left( t+1-v \right) \right\}]dv}{\int_{0}^{t} \exp\left\{ -Ʌ_{13}\left( v \right)-Ʌ_{14}\left( v \right)-Ʌ_{15}\left( v \right) \right\}.\lambda_{13}\left( v \right).[exp \left\{ -\Lambda_{35}\left( t-v \right) \right\}]dv}$$

$$\Pr_{t} \left\{ 5 | 3 \right\}=1-\Pr_{t} \{3|3\}$$

The probability of remaining in state (4) represents the conditional probability of staying in state (4) between time t and t+1 and death does not happen, given that the patient is in state (4) and alive at time t.

$$\Pr_{t} \left\{ 4 | 4 \right\}=\frac{\int_{0}^{t} \exp\left\{ -Ʌ_{13}\left( v \right)-Ʌ_{14}\left( v \right)-Ʌ_{15}\left( v \right) \right\}.\lambda_{14}\left( v \right).[exp \left\{ -\Lambda_{45}\left( t+1-v \right) \right\}]dv}{\int_{0}^{t} \exp\left\{ -Ʌ_{13}\left( v \right)-Ʌ_{14}\left( v \right)-Ʌ_{15}\left( v \right) \right\}.\lambda_{14}\left( v \right).[exp \left\{ -\Lambda_{45}\left( t-v \right) \right\}]dv}$$

$$\Pr_{t} \left\{ 5 | 4 \right\}=1-\Pr_{t} \{4|4\}$$

The probability of remaining in state (2) represents the conditional probability of staying in state (2) until t+1 and alive, given that the patient is in state (2) and alive at time t.

$$\Pr_{t} \left\{ 2 | 2 \right\}=\frac{exp\{-Ʌ_{25}\left( t+1 \right)\}}{exp\{-Ʌ_{25}\left( t \right)\}}$$

$$\Pr_{t} \left\{ 5 | 2 \right\}=1-\Pr_{t} \{2|2\}$$

The probability of remaining in state (6) represents the conditional probability of staying in state (6) until t+1 and alive, given that the patient is in state (6) and alive at time t.

$$\Pr_{t} \left\{ 6 | 6 \right\}=\frac{exp\{-Ʌ_{65}\left( t+1 \right)\}}{exp\{-Ʌ_{65}\left( t \right)\}}$$

$$\Pr_{t} \left\{ 5 | 6 \right\}=1-\Pr_{t} \{6|6\}$$

To focus our analysis on patient recurrence, we simplify the discrete-time Markov model by aggregating three states: "not-cured and no-persistent," "cured and persistent," and "persistent disease," into a single state labelled as "No recurrence." This simplification is motivated by the clinical importance of identifying recurrence, as clinicians primarily need to act upon detecting the recurrence type to initiate treatment. By combining states (1), (2), and (6) into "No recurrence," we streamline the model to emphasize the transitions that are most relevant for decision-making and treatment initiation. This aggregation is achieved using Bayesian rules, with the prior probabilities for these states being $p_{1}, p_{2}, \mathrm{and} p_{6}$. Based on this revised state structure, we construct a simplified Markov model and proceed with the optimization process accordingly.

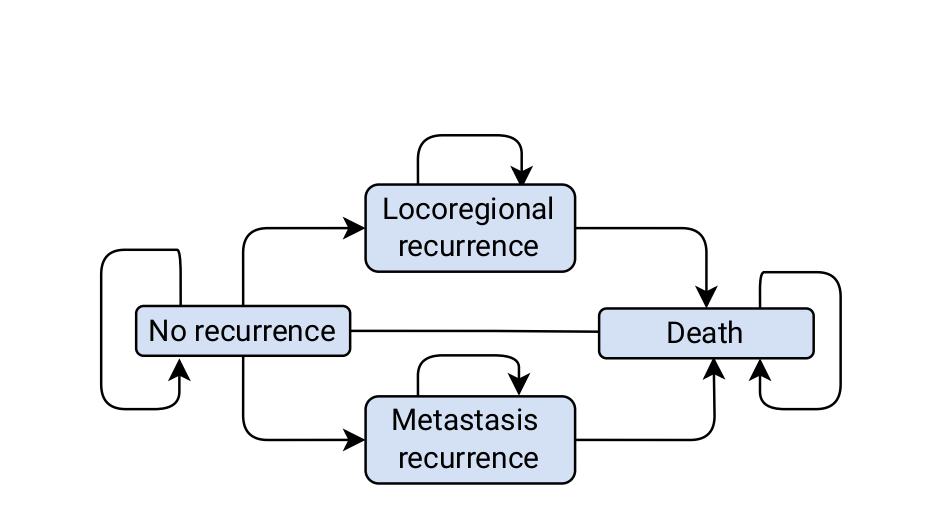

eFigure 2-Makov decision process, where states 1,2, and 6 are aggregated and named “No recurrence”

**Supplementary information 2. Optimization model and solution methodology**

In this section, we provide additional details related to the disease progression model and optimization model.

The model follows a hidden Markov model with the following components:

- **Time horizon**: the horizon length for the analysis is set to five years, with a cycle length of one month. The model begins with a patient whose cancer treatment has been completed.
- **State space**: The model consists of four states: "No recurrence" ($h_{n}),$ “Locoregional” $(h_{l})$, “Metastasis” ($h_{m})$, and the absorbing state "Death" ($D$).
- **Action space**: At each cycle, we decide whether to wait, perform a screening with a joint CT scan of the head and neck, or conduct a screening with a ctDNA blood-based test. Actions are represented by $a\in\{W,CT,DNA\}$.
- **Belief Space**: In the hidden Markov model, we lack precise knowledge of the patient's true state, as it is not directly observable. To address this uncertainty, we maintain a probability distribution over the hidden states $\{h_{n},h_{l},h_{m}\}$, referred to as the belief state $b$. The belief state represents our current estimate of the likelihood of the patient being in each hidden state, enabling us to make informed decisions based on this probabilistic understanding. We denote our belief space, the set of all possible beliefs, by $B$, defined as follows,

$$B=\left\{ b\in\left[ 0,1 \right]^{3}:\sum_{s\in\{h_{n},h_{l},h_{m}\}} b\left( s \right)=1 \right\}$$

- **Observation Set**: The set $O_{a}$ represents all potential observations that can result from taking an action $a\in\{W,CT,DNA\}$. In our framework:
  - Taking the action 𝑊 (wait) does not yield any immediate observations.
  - Taking the action CT (screening with a CT scan) can result in eight possible observations, as detailed in eTable 3. Any positive result from either CT-neck or CT-chest will be followed by a biopsy test.
  - Taking the action DNA (screening with a ctDNA test) can yield two possible observations: a positive or negative result.

eTable 3 Observation possibilities for screening with CT-scan

| **Observation** | **CT-neck** | **CT-chest** | **Biopsy-neck** | **Biopsy-chest** |
| --- | --- | --- | --- | --- |
| 1 | Negative | Negative | - | - |
| 2 | Negative | Positive | - | Positive |
| 3 | Negative | Positive | - | Negative |
| 4 | Positive | Negative | Positive | - |
| 5 | Positive | Negative | Negative | - |
| 6 | Positive | Positive | Positive | Negative |
| 7 | Positive | Positive | Negative | Positive |
| 8 | Positive | Positive | Negative | Negative |

- **Observation Probabilities**: the term $K\left( o | s \right)$ denotes the probability of observing $o\in O_{a}$ when the actual health state $s\in\{h_{n},h_{l},h_{m}\}$. For $o\in O_{w}$ the observation is null since no observation is generated when taking the action 𝑊 (wait). When performing screening actions CT-scan the observation probabilities are derived based on the sensitivity and specificity of the respective tests, as follows:

$$\alpha_{l}=Pr(CTneck=positive|s\neq locoregional)$$

$$\beta_{l}=Pr(CT-neck=negative|s=locoregional)$$

$$\alpha_{m}=Pr(CT-chest=positive|s\neq metastasis)$$

$$\beta_{m}=Pr(CT-chest=negative|s=metastasis)$$

$\bar{\alpha_{l}}=1-\alpha_{l}=specificity of CT-neck$

$$\bar{\beta_{l}}=1-\beta_{l}=sensitivity of CT-neck$$

$$\bar{\alpha}_{m}=1-\alpha_{m}=specificity of CT-chest$$

$$\bar{\beta}_{m}=1-\beta_{m}=sensitivity of CT-chest$$

The observation probability for screening with the CT-scan of both head and neck cancer is following eTable 4.

eTable 4 Observation probabilities for all possible CT-scan/biopsy result

| observation/state | No Cancer | Locoregional | Metastasis |
| --- | --- | --- | --- |
| 1 | $\bar{\alpha_{l}}\bar{\alpha}_{m}$ | $\beta_{l}\bar{\alpha}_{m}$ | $\bar{\alpha}_{l}\beta_{m}$ |
| 2 | 0 | 0 | $\bar{\alpha_{l}}\bar{\beta}_{m}$ |
| 3 | $\bar{\alpha}_{l}\alpha_{m}$ | $\beta_{l}\alpha_{m}$ | 0 |
| 4 | 0 | $\bar{\beta_{l}}\bar{\alpha}_{m}$ | 0 |
| 5 | $\alpha_{l}\bar{\alpha}_{m}$ | 0 | $\alpha_{l}\beta_{m}$ |
| 6 | 0 | $\bar{\beta}_{l}\alpha_{m}$ | 0 |
| 7 | 0 | 0 | $\alpha_{l}\bar{\beta}_{m}$ |
| 8 | $\alpha_{l}\alpha_{m}$ | 0 | 0 |

The observation probabilities for screening with ctDNA is as follows:

$$\alpha_{\mathrm{DNA}}=Pr(ctDNA=positive|s=No Cancer)$$

$$\beta_{\mathrm{DNA}}=Pr(ctDNA=negative|s \neq No Cancer)$$

$\bar{\alpha}_{DNA}=1-\alpha_{\mathrm{DNA}}=$specificity of ctDNA

$\bar{\beta}_{DNA}=1-\beta_{\mathrm{DNA}}=$sensitivity of ctDNA

- **Transition Probabilities**: We denote the transition probabilities between the core states by $P_{t}(s^{'}|s,a)$ and transition probabilities between the belief states by $P_{t}(b^{'}|b,a)$ for action $a\in\{W,CT,DNA\}$. The death and survival probabilities in period t is denoted by $d_{t}$ and $\bar{d}_{t}$, respectively.
- **Belief Update**: The model engages in decision-making, where new observations $o\in O_{a}$are obtained as a result of taking any screening action $a\in\{CT,DNA\}$. When a new observation is received, the belief state is updated according to the following equation:

$b\leftarrow\tau_{1}\left[ b,o,a \right]\left( s^{'} \right)=\frac{b\left( s^{'} \right)k(o|s^{'})}{\sum_{s\in\{h_{n},h_{l},h_{m}\}} b\left( s \right)k(o|s)},$ $\forall s^{'}\in\left\{ h_{n},h_{l},h_{m} \right\}.$

Where $\tau_{1}[b,a,o](s^{'})$ is the posterior probability of being in hidden state $s^{'}\in\{h_{n},h_{l},h_{m}\}$ after taking screening action $a$ and observing $o\in O_{a}$.

In the case that cancer is not detected, or the wait action is taken, state transition occurs, and the belief state undergoes the following update:

$b\leftarrow\tau_{2}\left[ b \right]\left( s^{'} \right)=\sum_{s\in\{h_{n},h_{l},h_{m}\}} b\left( s \right)P_{t}(s^{'}|s)$, $\forall s^{'}\in\{h_{n},h_{l},h_{m}\}$.

Where $\tau_{2}[b](s^{'})$ is the posterior probability of being in hidden state $s^{'}\in\{h_{n},h_{l},h_{m}\}$ after transition between states happens. These equations provide a clear representation of how the belief state evolves in response to both observations and state transitions, ensuring that the model accounts for changing information and circumstances and it forms the basis for determining optimal actions in subsequent decision-making steps.

- **Rewards**: At each time epoch, there are two types of rewards that can occur: detection delay, and the cost of screening. The computation of each reward is explained as follows:

$d(s,a)=$expected delay of detection between two-time epochs when the true health state is s and action a is taken as follows.

- If $a=wait, s\in\{h_{l},h_{m}\}$, $d\left( s,a \right)=1$
- If the action $a$ is to screen with a joint CT scan of the neck and chest, and the patient is in state $s\in\{h_{l}\}$, detection might be delayed due to the test's accuracy limitations. The test may still yield a negative result even when the patient is in the locoregional state, with probability $\beta_{l}$. Therefore, the detection delay is given by $d\left( s,a \right)=\beta_{l}$.
- If the action $a$ is to screen with a joint CT scan of the neck and chest, and the patient is in state $s\in\{h_{m}\}$, detection might be delayed due to the test's accuracy limitations. The test may still yield a negative result even when the patient is in the metastasis state, with probability$\beta_{m}$. Therefore, the detection delay is given by $d\left( s,a \right)=\beta_{m}$.
- If the action $a$ is to screen with a ctDNA test, and the patient is in state $s\in\{h_{l},h_{m}\}$, detection will be delayed by one month, as treatment cannot begin immediately. Therefore, the detection delay is given by $d\left( s,a \right)=1$.

$C(s,a)=$expected cost of screening when the true health state is s and action a is taken as follows. We represent cost of biopsy with $C_{biop}$, cost of CT-scan with $C_{CT}$, and cost of ctDNA with $C_{DNA}$

- If $a=wait, d\left( s,a \right)=0$
- If the action $a$ is to screen with a joint CT scan of the neck and chest, and the patient is in state $s\in\{h_{n}\}$, due to the false positive results, there is a chance that the patient is sent for the biopsy test. Therefore, the expected cost is as follows:

$$C\left( s,a \right)=C_{CT}+\left[ \alpha_{l}\left( 1-\alpha_{m} \right)+2\alpha_{l}\alpha_{m}+\left( 1-\alpha_{l} \right)\alpha_{m} \right].C_{biop}$$

- If the action $a$ is to screen with a joint CT scan of the neck and chest, and the patient is in state $s\in\{h_{l}\}$, the patient is referred for a biopsy if the neck CT scan is positive or due to false positive risk, if the chest CT scan is positive. The expected cost in this case is as follows:

$$C\left( s,a \right)=C_{CT}+[\bar{\beta}_{l}.\bar{\alpha}_{m}+2\bar{\beta}_{l}.\alpha_{m}].C_{biop}$$

- If the action $a$ is to screen with a joint CT scan of the neck and chest, and the patient is in state $s\in\{h_{m}\}$, the patient is referred for a biopsy if the chest CT scan is positive or due to false positive risk, if the neck CT scan is positive. The expected cost in this case is as follows:

$$C\left( s,a \right)=C_{CT}+[\bar{\beta}_{m}.\bar{\alpha}_{l}+2\bar{\beta}_{m}.\alpha_{l}].C_{biop}$$

- If $a=screen with ctDNA, s\in\{h_{n},h_{l},h_{m}\}$

$$C\left( s,a \right)=C_{DNA}$$

We define $r\left( s,a \right)=\lambda.d\left( s,a \right)+c(s,a)$, where λ is a factor that converts $d(s,a)$ into its monetary equivalent. By adjusting and fine-tuning the value of 𝜆, we can control the trade-off between detection delay and cost, allowing us to achieve any desired target detection delay. To compute the reward for a belief state, we use the following formula:

$r\left( b,a \right)=\sum_{s\in\{h_{n},h_{l}, h_{m}\}} b\left( s \right).r(s,a)$,

where $b(s)$ is the belief (probability) of being in state $s$. This formula calculates the expected reward for a given belief state and action.

- **Optimality Equations**: Since we impose the restriction that two consecutive CT-scan tests cannot be conducted with a time difference of less than three months, we define $l$ as the remaining time until a CT-scan becomes available. The variable $l$ can take values from {0,1,2}, where $l=0$ indicates that a CT-scan decision can be incorporated, while $l>0$ restricts the available actions to either waiting or conducting a ctDNA test. To account for this restriction, we define $v_{t}(b,l)$ as the value function representing the expected utility at belief state $b$ with the remaining time $l$ until a CT-scan becomes available at time $t$. When $t=T$ (the final time step), the value function is governed by the following equations:

$$v_{T}\left( b,l \right)=\min_{a\in\{w,c_{T},c_{d}\}} \{r_{T}(b,a)\}, \forall b\in B, l\in\{0,1,2\}$$

For $l=0 , b\in B,$ and $t<T$:

$$v_{t}\left( b,l \right)=\min_{a\in\left\{ W,CT, DNA \right\}} \{r_{t}\left( b,a \right)+\gamma.\bar{d_{t}}.\left[ \sum_{b^{'},a\in\left\{ W, DNA \right\}} \Pr_{t} \left\{ b^{'} | b,a \right\}.v_{t+1}\left( b^{'},0 \right)+P\left\{ b^{'} | b,CT \right\}.v_{t+1}\left( b^{'},2 \right) \right]\}$$

For $l>0, b\in B$ and $t<T$, the CT scan is unavailable due to the three-month restriction. The value function is defined as:

$v_{t}\left( b,l \right)=\min_{a\in\left\{ W, DNA \right\}} \{r_{t}\left( b,a \right)+\gamma.\bar{d_{t}}.\sum_{b^{'},a} \Pr_{t} \left\{ b^{'} | b,a \right\}.v_{t+1}\left( b^{'},l-1 \right)\}$,

where $v_{t+1}\left( b^{'},l-1 \right)$ represents the value function at the next time step with $l$ decremented by one. As $l$ decreases by one at each step, it eventually reaches zero, at which point the CT scan becomes available again.

**Solution Methodology**: To solve the above equations and derive an implementable policy, we employ the following steps:

- *Policy Optimization Using Backward Induction:*

The first step involves solving the optimality equations using the backward induction method. This approach enables us to determine the optimal policy for each belief state while accounting for the dynamic progression of the problem. The backward induction process iterates backward through time, ensuring that the decisions minimize the expected rewards given the belief states.

However, implementing these policies in real-world settings is challenging. The original optimal policy is defined as a function of the belief state, which represents the probability distribution over the hidden states. Keeping track of and updating the belief state in clinical practice requires complex calculations and continuous monitoring, which may not be practical. These challenges necessitate the development of an easy-to-implement policy that preserves the key insights from the optimal solution while simplifying its application.

- *Determining Testing Intervals Through Simulation:*

To address the limitations of the original policy, we use the optimal policies derived from the backward induction step as inputs to a simulation engine. The simulation replicates patient disease trajectories over time under the surveillance strategy. By analysing the progression of disease states and the policy behaviour, we identify the intervals between consecutive ctDNA tests. These intervals are extracted as a simplified representation of the optimal policy, ensuring it is easier to implement in clinical practice. From the patient disease trajectories and the optimized policies, we observed that the policy can effectively be represented as the time intervals between ctDNA tests. This time-based structure simplifies decision-making while retaining the performance benefits of the optimization approach. In eTable 6, we present the tabular format of the ctDNA schedule for all sub-populations.

eTable 6	ctDNA schedule for different target recurrence detection delay for all sub-populations

| Sub-population | ctDNA schedule (month) for target recurrence detection delay | | |
| --- | --- | --- | --- |
|  | 9 months | 6 months | 3 months |
| Stage 1, non-smoker | 10, 21 | 9, 16, 23, 35 | 3, 5, 7, 10, 14, 20, 30 |
| Stage 1, ever-smoker | 15 | 11, 20, 25 | 6, 10, 13, 17, 22, 29 |
| Stage 2, non-smoker | 10, 23 | 8, 15, 22, 27 | 5, 8, 12, 15, 18, 22, 27, 34 |
| Stage 2, ever-smoker | 16 | 8, 17, 26 | 6, 10, 15, 20, 25, 29, 34 |
| Stage 3, non-smoker | 8, 16 | 5, 9, 15, 21 | 4, 7, 10, 14, 20, 25, 28, 32 |
| Stage 3, ever-smoker | 15 | 8, 17, 24 | 4, 6, 9, 13, 17, 22, 28, 36 |
| Unstratified population | 11 | 9, 20, 30 | 5, 8, 12, 18, 21, 26, 32 |

**Supplementary information 3. Population aggregation method**

To create sub-populations based on different cancer stages and smoking statuses, we apply a Bayesian rule, utilizing the prior distribution of patients defined by their covariates as detailed in eTable 5. Let $q_{i}$ represent the distribution of patients with covariate $i$, $\Pr_{t}^{i} \{s^{'}|s\}$ denote the transition probability from state $s$ to $s'$ at time $t$ for patients with covariate $i$, and $\pi_{t}^{i}(s)$ be the occupancy probability of being in state $s$ at time $t$ for patients with covariate $i$. To construct a sub-population that is invariant with respect to the $|I|$ covariates and variant to cancer stage and smoking status ($j\in J$), we aggregate over the covariates using the following formula:

- For $t=1$, we compute the expected transition probabilities between states for $|I|$ covariates, where the weights are given by $q_{i}$:

$${\Pr_{t}}^{j} \{s^{'}|s\}=\sum_{i}^{\left| I \right|} q_{i}.\Pr_{t}^{i} \{s^{'}|s\}, s,s^{'}\in\{h_{n},h_{l},h_{m},D\}$$

- For $t>1,$ we use Bayes' law to compute the transition probabilities, which determine the weighted probability of transitioning from state $s'$ and $s$ given that the patient is in state $s$ at $t-1$, the transition probability is derived as follows:

${\Pr_{t}}^{j} \left\{ s^{'} | s \right\}=\frac{\sum_{i}^{\left| I \right|} q_{i}\pi_{t-1}^{i}\left( s \right)\Pr_{t}^{i} \{s^{'}|s\}}{\sum_{i}^{\left| I \right|} q_{i}\pi_{t-1}^{i}\left( s \right)}, s,s^{'}\in\{h_{n},h_{l},h_{m},D\}$,

Here, $\pi_{t-1}^{i}(s)$ represents the occupancy probability of being in state $s$ at time $t-1$ for covariate $i$. This probability is obtained by successively multiplying the transition matrices for sub-population $i$ up to time $t-1$.

In eFigure 3 and 4, we present the transition probabilities from no-recurrence state and occupancy probabilities over time for all six sub-populations.

eTable 5 Distribution of the patient’s characteristics in the population

| Characteristics | Median (IQR) | Source |
| --- | --- | --- |
| Age at diagnosis, y | 58 (52-64.4) | [1] |
|  | Percentage |  |
| Sex |  |  |
| Male | 85.1% | [2] |
| Female | 14.9% | [2] |
| Anemia |  |  |
| No | 78.1% | [1] |
| Yes | 21.9% | [1] |
| ACE27 comorbidity |  |  |
| None | 31% | [1] |
| Mild | 37.2% | [1] |
| Moderate | 19.4% | [1] |
| Severe | 12.4% | [1] |
| Smoking status |  |  |
| Never | 33.2% | [1] |
| Former | 34.7% | [1] |
| Current | 32.1% | [1] |
| T classification (AJCC 8^th^ edition) |  |  |
| 1 | 26.4% | [2] |
| 2 | 37.5% | [2] |
| 3 | 21.6% | [2] |
| 4 | 14.4% | [2] |
| N classification (AJCC 8^th^ edition) |  |  |
| 0/1 | 70% | [2] |
| 2 | 23% | [2] |
| 3 | 7% | [2] |

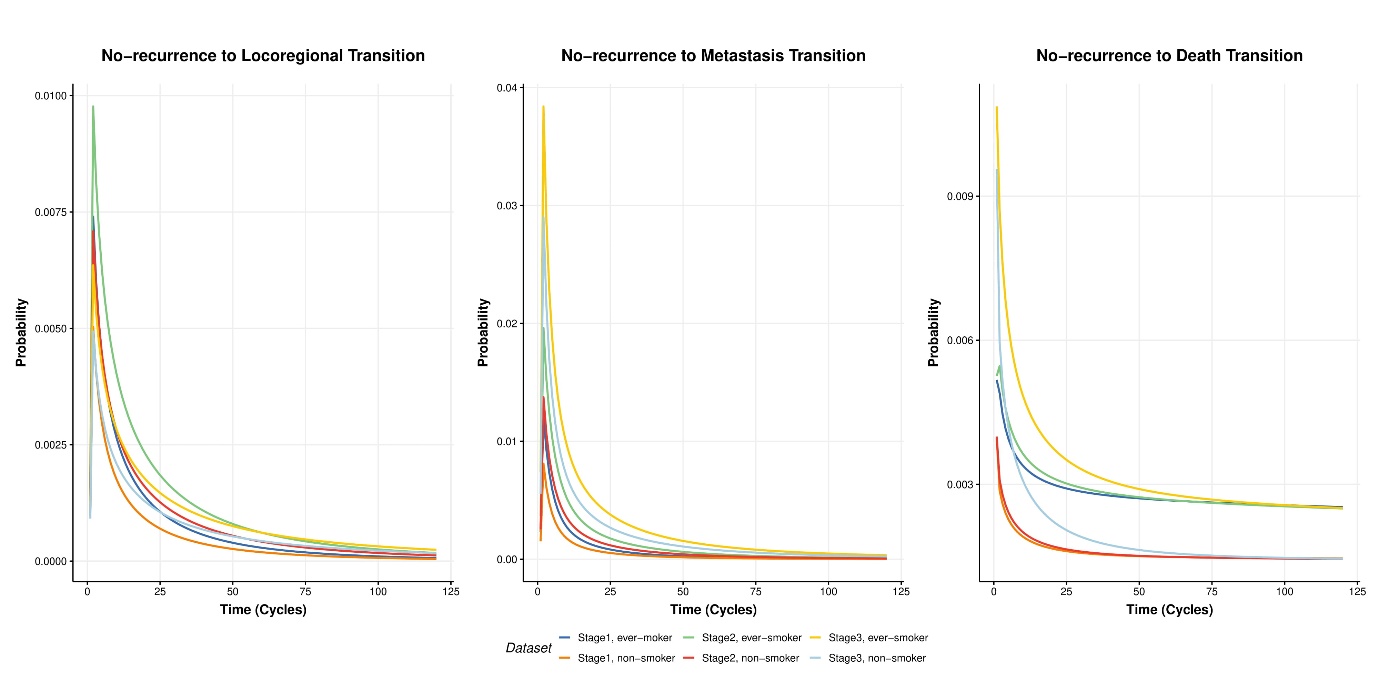

eFigure 3 Transition Probabilities from No-recurrence state for six sub-populations.

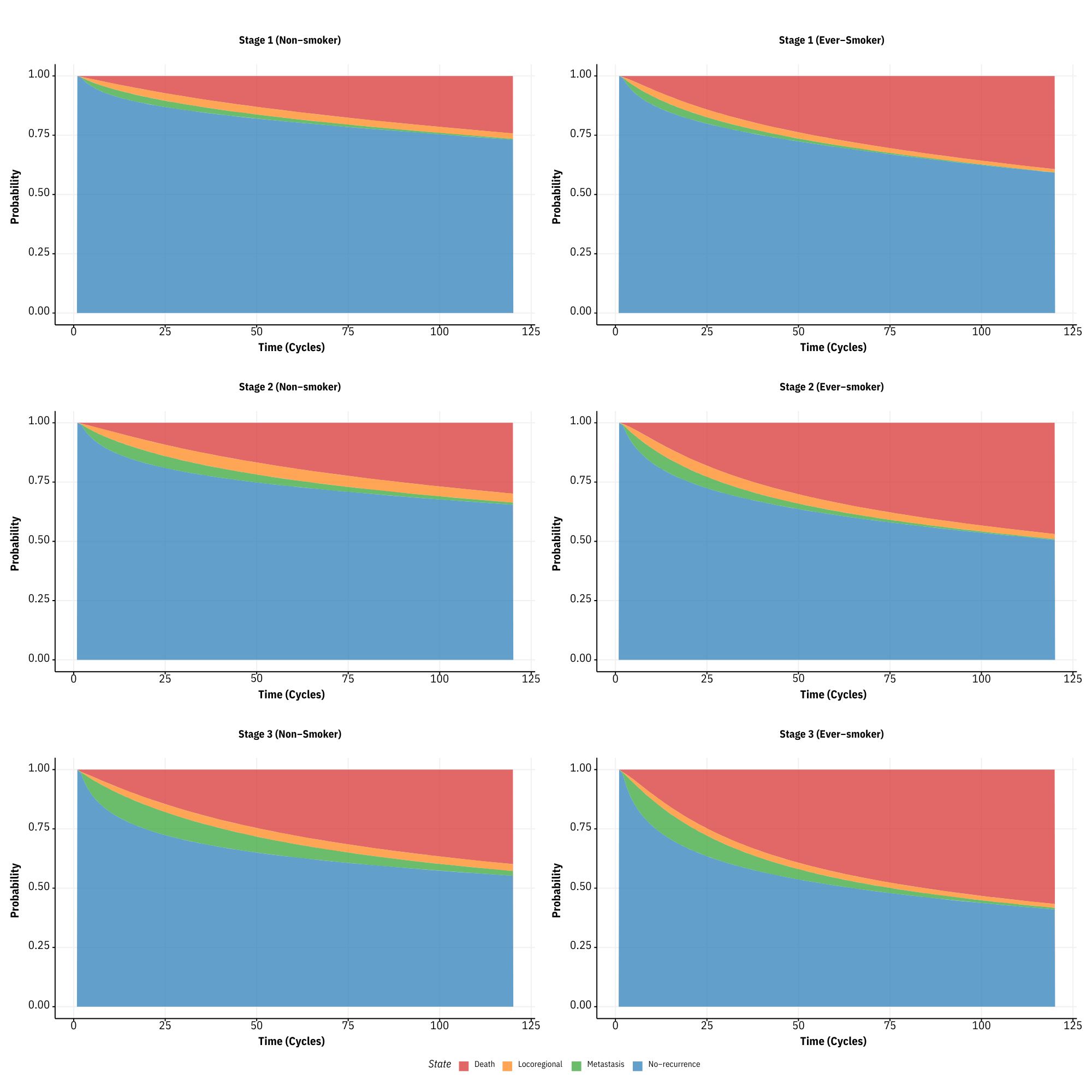

eFigure 4 Occupancy Probabilities over time for six sub-populations
